# Endothelial injury and acute-phase inflammatory mediators converge to drive dengue severity

**DOI:** 10.64898/2026.03.18.26348673

**Authors:** Abdul R. Anshad, Muthuvel Atchaya, Shanmugam Saravanan, Amudhan Murugesan, Pachamuthu Balakrishnan, Sivadoss Raju, Yong K. Yong, Marie Larsson, Esaki M. Shankar

**Author notes:** **Correspondence:** Esaki M Shankar,; Marie Larsson.

## Abstract

**Introduction:** Severe dengue infection is characterized by endothelial injury and systemic inflammatory complications. To better understand the mechanisms underlying disease severity, we investigated a broad panel of circulating inflammatory and endothelial mediators in patients with clinical dengue infection.

**Methods:** A prospective cross-sectional case–control study was carried out involving 111 dengue patients and 42 healthy controls. Among the dengue cases, 85 were identified as primary, while 26 were classified as secondary dengue infections. Serum levels of endothelial markers (Ang-2, CXCL10, MCP1, TRAIL), acute-phase and liver dysfunction and acute-phase markers (CRP, galectin 3, and serum amyloid protein), systemic inflammatory mediators (MIF, TNF-α, IL-1β), mast cell-derived proteases (chymase, tryptase), and tissue repair markers HGF, IL-10, IL-1Ra) were quantified using ELISA and Luminex multiplex assays. Correlations among serum analytes, severity indicators, and haematological markers were also explored

**Results:** Several biomarkers, Ang-2, CXCL10, TRAIL, CRP, MIF, IL-1Ra, TNF-α, and chymase showed differential expression across severity groups, indicating coordinated endothelial and inflammatory activation. Stratification of patients with primary-secondary dengue also followed a similar pattern except IL-1β, which had significant differential expression across the cohorts. Ang-2 showed strong positive correlations with markers of hepatic dysfunction, including ALT, AST, and bilirubin, suggesting a link between endothelial injury and liver involvement.

**Conclusions:** Severe dengue is driven by the coordinated activation of endothelial dysfunction, acute-phase responses, mast cell mediators, and counter-regulatory pathways. These processes collectively contribute to vascular leakage and organ injury, reinforcing the value of biomarkers such as Ang-2, CXCL10, CRP, and chymase for severity assessment.

## Introduction

Dengue virus (DENV), a member of *the Flaviviridae family,* transmitted by female *Aedes aegypti* and *Aedes albopictus, infects* ∼400 million people annually and remains hyperendemic in many tropical and sub-tropical regions (Anshad et al., 2026; Halstead & Wilder-Smith, 2019; Nanaware et al., 2021). With expanding mosquito geographical distribution, driven in part by anthropogenic activities and global temperature changes, dengue distribution appears to expand beyond endemic regions where it is less reported otherwise (Meena et al., 2020; Paz-Bailey et al., 2024). The increasing incidence of dengue, coupled with diverse clinical presentations across serotypes and primary versus secondary infections, underscores the need for improved diagnostic and prognostic tools, especially to predict dengue severity (Narvaez et al., 2025). Based on the clinical features and disease progression, dengue is classified into three primary categories: dengue without warning signs (DWS-), dengue with warning signs (DWS+), and severe dengue (SD) (Giang et al., 2020). Dengue disease progression is influenced by a combination of immune dysregulation and direct viral effects, and the specific pathomechanisms remain inscrutable (Khanam et al., 2022). Clinical presentation ranges from subclinical/asymptomatic dengue to fatal disease encompassing severe thrombocytopenia and hypovolemic shock, as well as multi-organ impairment (Imad et al., 2020). Severe complications of the multi-organ impairment could culminate in poor prognosis e.g., vascular derangement, renal dysfunction, dengue meningitis, and hepatic encephalopathy (Goswami et al., 2012; Vaidya & Salvi, 2023).

Endothelial dysfunction represents the central axis of dengue pathogenesis and is a key driver of severe disease contributing to thrombocytopenia and dengue-associated shock (Castilho et al., 2020; Dalrymple & Mackow, 2012). Mediators such as angiopoietin 2 (Ang-2), CXCL10/IP-10, CCL2/MCP-1, TNFα, IL-1β, and TRAIL (TNF-related apoptosis inducing ligand)/TNSF-10 reflect endothelial activation, leukocyte recruitment, and apoptosis, which collectively contribute to increased vascular permeability and capillary leakage by destabilising the endothelial junction (Cipitelli et al., 2022; Mishra et al., 2024). Hepatic involvement represents another prominent feature of dengue infection, ranging from mild transaminitis to acute liver dysfunction (Samanta, 2015). The DENV induced hepatic-immune modulation can be measured using acute-phase proteins, C-reactive protein (CRP), and serum amyloid protein (SAP), as well as immune markers, including galectin 3, macrophage migratory inhibitory factor (MIF), IL-10, TNFα, IL-1β, and CXCL10. Together these markers indicate both hepatic dysfunction and the broader systemic dengue disease burden (Brasier et al., 2012; Conroy et al., 2015; Lai et al., 2020; Mukherjee et al., 2022).

Aside from hepatic involvment, pulmonary and renal disorders are also increasingly recognised in clinical dengue infection (Gurugama et al., 2018; Keshav et al., 2024). Pulmonary disorders including acute lung injury or acute respiratory distress syndrome (ARDS) are closely associated with increased endothelial permeability (Mehmood et al., 2023). Further, DENV-mediated mast cell activation appears to be a determinant of clinical dengue as measured by the levels of chymase, tryptase, and TNF-α, which are indicative of capillary permeability and acute-phase inflammatory responses. In contrast, the tissue repair as well as counter-regulatory pathways in clinical dengue is often reflected by the relative concentrations of hepatocyte growth factor (HGF) and IL-1 receptor antagonist (IL-1RA), which affirms the host’s attempt to resolve inflammation during the recovery phase to restore endothelial integrity. In this study, we investigated a broad range of systemic mediators of inflammation and endothelial dysfunction to underpin their potential associations with disease severity in dengue infection.

## Materials and Methods

### Ethics approval

The study was reviewed and approved by the Institutional Ethical Committee (IEC) of the SMCH, Chennai (Ref. No. 114/03/2024/Faculty/SRB/SMCH) and TMC, Theni (Ref. No. 2300/IEC/2024-26), and was carried out in accordance with the Guidelines of the International Conference on Harmonization Guidelines and the Declaration of Helsinki (1964). All the human subjects were adults and provided written consents to participate in the study. Blood specimens were collected in BD Vacutainer tubes (Becton Dickinson, Franklin Lakes, NJ, USA) by a trained phlebotomist and stored at –80°C until use in the experiments in 100 µL aliquots to avoid multiple freeze-thaw cycles.

### Study population and clinical classification

DENV-infected individuals were recruited initially based on clinical symptoms, and were further confirmed by dengue-NS1 antigen or anti-dengue IgM ELISA (Anshad et al., 2026; Sánchez et al., 2024). The NS1 antigen test was performed at the time of blood collection by the clinicians using a commercial NS1 ELISA kit (cat. no. 01PE40, Abbott, Chicago, IL, USA). PanBio dengue IgM (cat. no. 01PE20, Abbot, USA) and PanBio dengue IgG (cat. No. 01PE10, Abbot, USA) kits were used to classify the participants/ patients into primary and secondary dengue. Samples with positivity in both ELISAs with an IgM/IgG of ≤1.1 were considered secondary dengue, whereas IgM^+^IgG^+^ samples with an IgM/IgG ratio >1.1 or IgM^+^IgG^−^ were categorized as primary dengue.

### Luminex multiplex assay

Serum concentrations of chemokines and cytokines were quantified by a commercial Bio-Plex Luminex multiplex assay (Bio-Rad Laboratories, Hercules, CA, USA) with the Human Luminex Discovery 9-plex (cat. no. LXSAHM-09, R&D Systems, Minneapolis, USA). The analytes measured were CCL2/JE/MCP-1 (Monocyte chemoattractant protein-1), HGF (hepatocyte growth factor), IL-1Ra/IL-1F3, MIF (macrophage migration inhibitory factor), TRAIL (TNF-related apoptosis-inducing ligand)/TNFSF-10, CXCL10/IP-10/CRG-2/IL-1β/IL-1F2, IL-10, and TNF-α (Tumor necrosis factor alpha). Frozen serum stored in batches were thawed at 4°C before diluting at a 1:4 ratio using sample diluent and analyzed on a Bio-Plex 200 system according to the manufacturer’s instructions.

### Enzyme-linked immunoassays

Commercial ELISAs were employed for measuring circulating liver dysfunction and acute-phase mediators such as serum amyloid P (SAP) (cat. no. ab246537, Abcam, Cambridge, UK; sensitivity 181pg/ml), nephritic marker galectin-3 (cat. no. ab269555, Abcam, Cambridge, UK; sensitivity 13.3pg/ml), and C-reactive protein (CRP) (cat. no. DCRP00B, Biotechne, Minneapolis, USA; sensitivity 0.022ng/ml), were quantified as per the manufacturer’s instructions. The endothelial activation marker, angiopoietin-2 (Ang-2) was also measured using a commercial ELISA (cat. no. DANG20, Biotechne, Minneapolis, USA sensitivity 21.3pg/ml). Besides, the mast cell-specific proteases, tryptase (cat. no. ab314709, Abcam, Cambridge, UK; sensitivity 2ng/ml) and chymase (cat. no. CSB-E13757h, Cusabio, Houston, USA; sensitivity 0.039ng/ml), were also measured by commercial ELISA.

### Statistical analysis

Data were analysed using Kruskal–Wallis’s test followed by Mann– Whitney U-tests post-hoc tests to compare levels of each biomarker across the DWS-, DWS+, and SD groups as well as the primary and secondary dengue. Logistic regression models were constructed to assess the associations between specific biomarkers and the risk of severe disease. Correlation analysis was performed to determine the relationship between immune markers and clinical severity scores. Statistical significance was set at a P value <0.05 for all the analyses. This comprehensive methodology allowed for the examination of immune dysregulation, vascular involvement, and renal impairment markers in dengue patients. By profiling these markers across different severity levels, we aimed to elucidate the molecular and immunological drivers of severe dengue, with a focus on identifying biomarkers that could guide early diagnosis and targeted intervention.

## Results

### Clinico-demographic data of participants recruited in the dengue investigation

In this prospective cross-sectional case-control study, we recruited 153 participants, which included 111 dengue patients encompassing primary (n=85) and secondary (n=26) infections, and healthy controls (n=42). In the dengue cohort, 35 participants were positive for anti-DENV IgG, while 103 were positive for anti-DENV IgM. Twenty-four subjects were positive for NS1, of which 18 tested positive for both IgM and NS1 (**Table 1**). All participants in the study were adults, aged over 18 years. The workflow of the investigation is illustrated in **Figure 1**.

**Figure 1.**
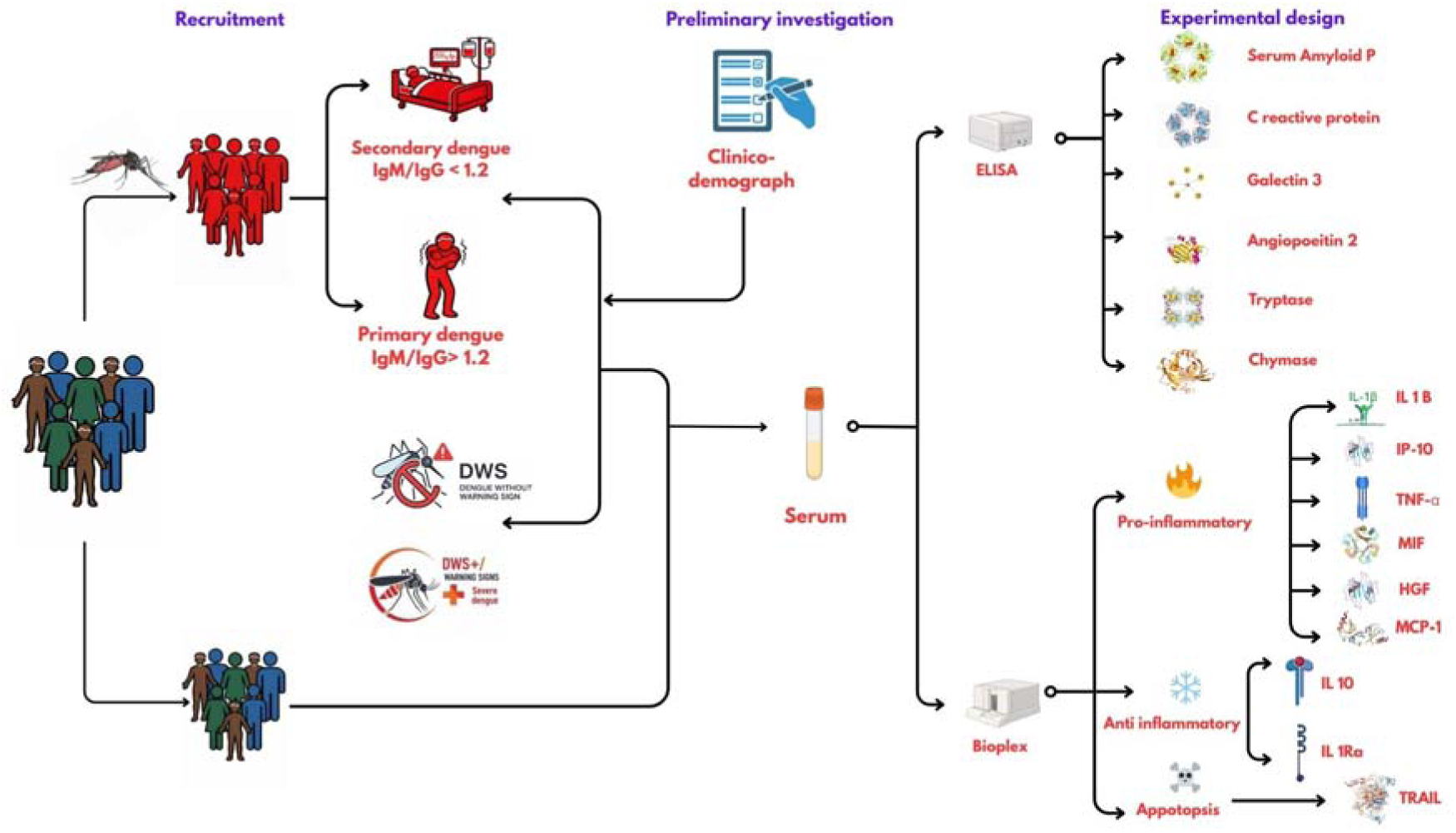
Graphical abstract of the cross-sectional case-control investigation of organ dysfunction markers as well as cytokines. The cohort was divided in to dengue with warning signs, dengue without warning signs, severe dengue which also stratified in another cohort of primary and secondary dengue based on WHO 2007 classification. The serum samples were collected from adult patients/volunteers with proper consent and were measured using ELISA as well as Bioplex Luminex cytokine array systems.

**Table 1:**
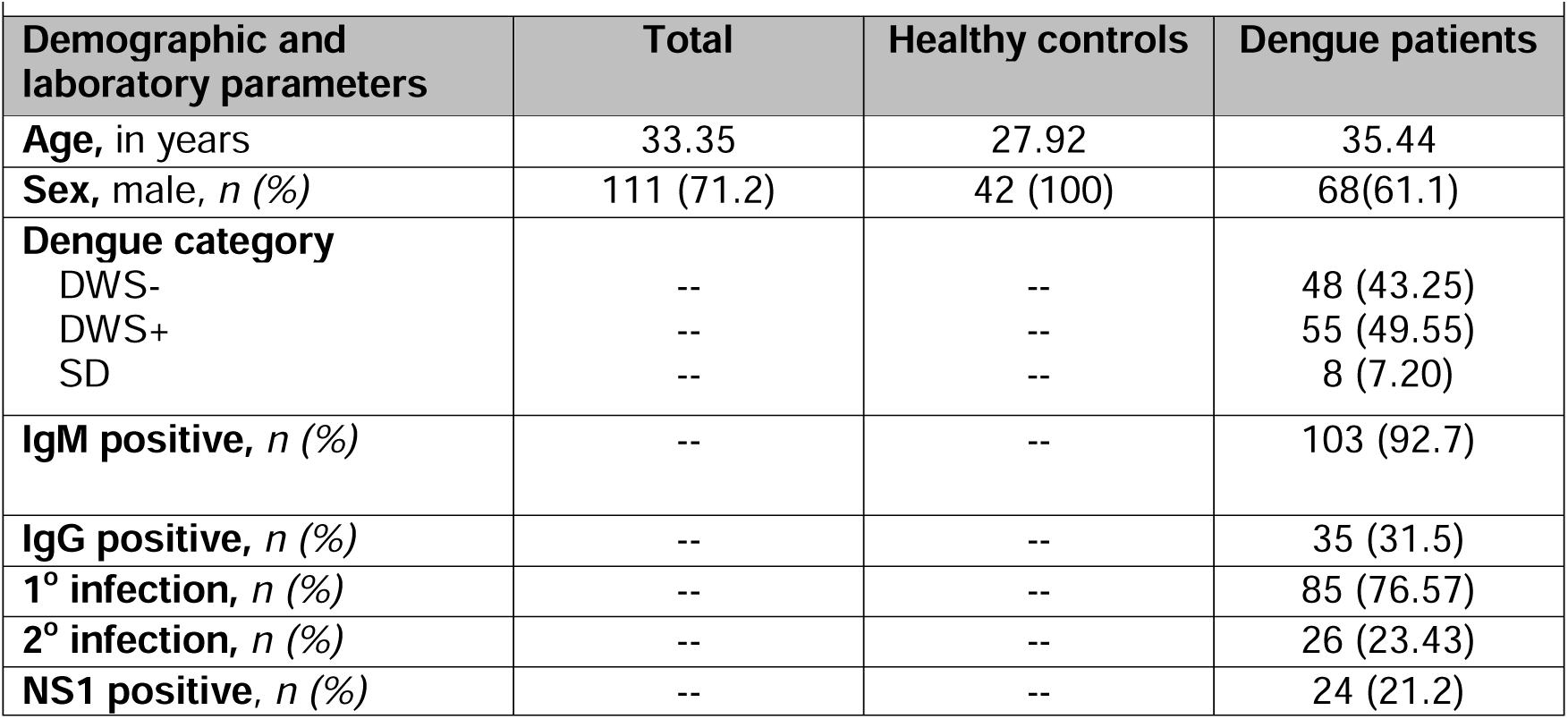
Cohort characteristics for demography and laboratory characters at admission. Table 1. **Patients’ characteristics for demography, laboratory characteristics at admission**. Comparison of demographic and clinical characteristics between dengue patients and healthy control. Percentages are given in the bracket. **Footnotes:** DWS-, Dengue without warning signs; DWS+, Dengue with warning signs; SD, Severe dengue; MBL, 1° infection, Primary infection; 2° infection, secondary infection; NS1, Non-structural protein 1.

### Mediators linked to endothelial dysfunction and capillary permeability markers were significantly elevated in dengue with warning signs and severe dengue

Mediators of endothelial dysfunction and vascular permeability were significantly elevated in DWS+ and in those with severe disease. CXCL10 levels were markedly higher in all dengue groups relative to the healthy controls, with highest increase observed among DWS+ (P<0.0001) and SD (P<0.0001) groups (**Figure 2**). Ang-2 also showed a progressive increase, with the highest levels observed in the SD (P<0.05) as well as DWS+ (P<0.05) groups than in the DWS-cohort (P<0.001), while remaining lower in the healthy cohort (**Figure 3**). CCL2/MCP-1 (**Figure 2**) levels were moderately elevated in the dengue cohorts, although not significantly so. In contrast, TRAIL/TNFSF-10 levels (**Figure 2**) were increased in dengue patients with a higher elevation observed across both DWS+ (P<0.05) and SD (P<0.05) patients than the DWS-patients (P<0.01). Overall, these findings indicated that inflammatory mediators associated with endothelial activation and capillary permeability were markedly elevated in severe dengue infection.

**Figure 2.**
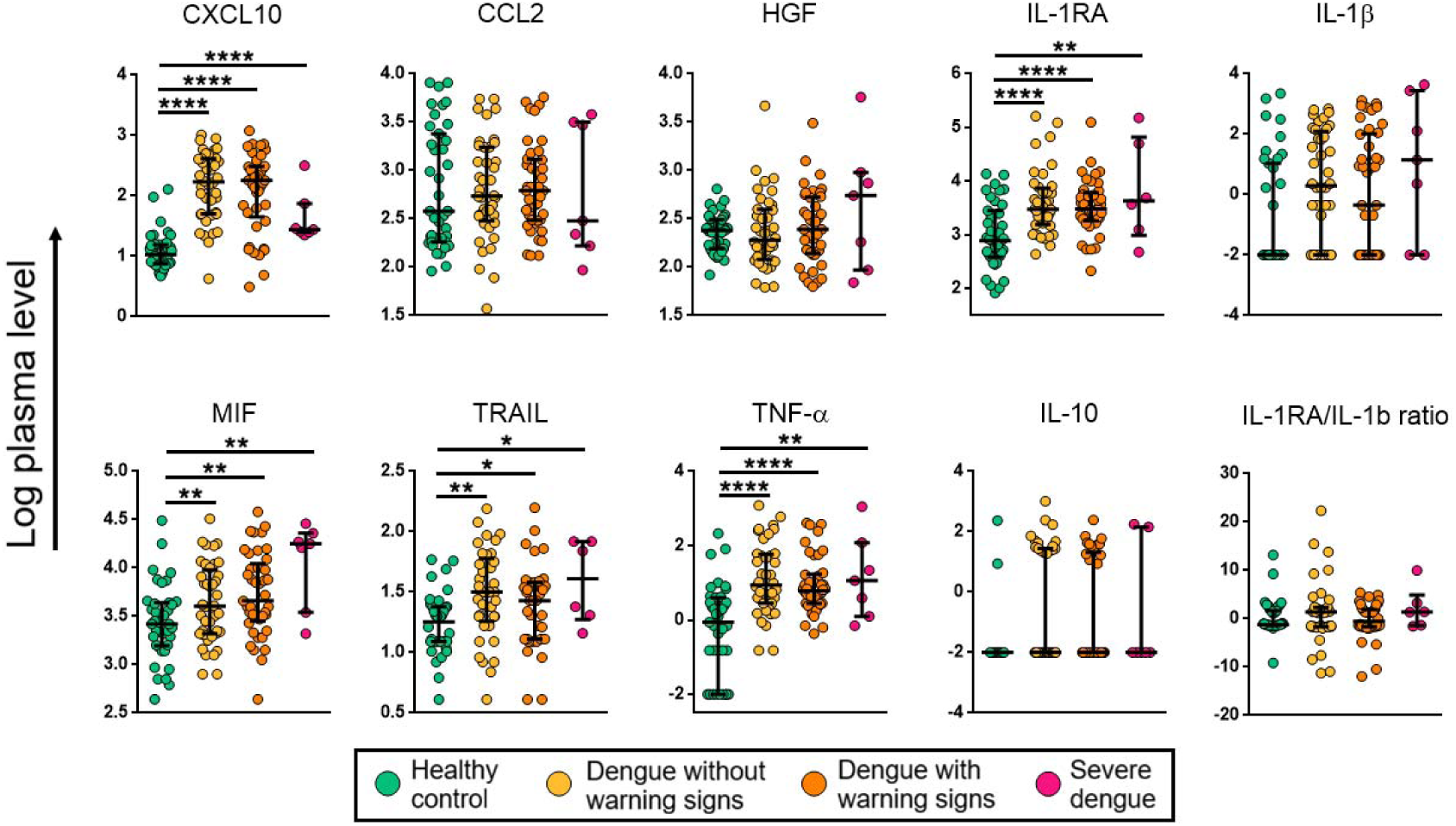
Comparison of serum levels of cytokines in healthy controls, dengue without warning signs, dengue with warning signs and severe dengue compared with healthy controls. Levels of biomarkers were compared across the four study groups by the Kruskal–Wallis test. Post hoc Mann–Whitney U-tests were subsequently performed for those complement biomarkers with a Kruskal–Wallis P-value of <0.05. P-value <0.05 (significant); *, <0.05; **, <0.01; ***, <0.001, ****, <0.0001.

**Figure 3.**
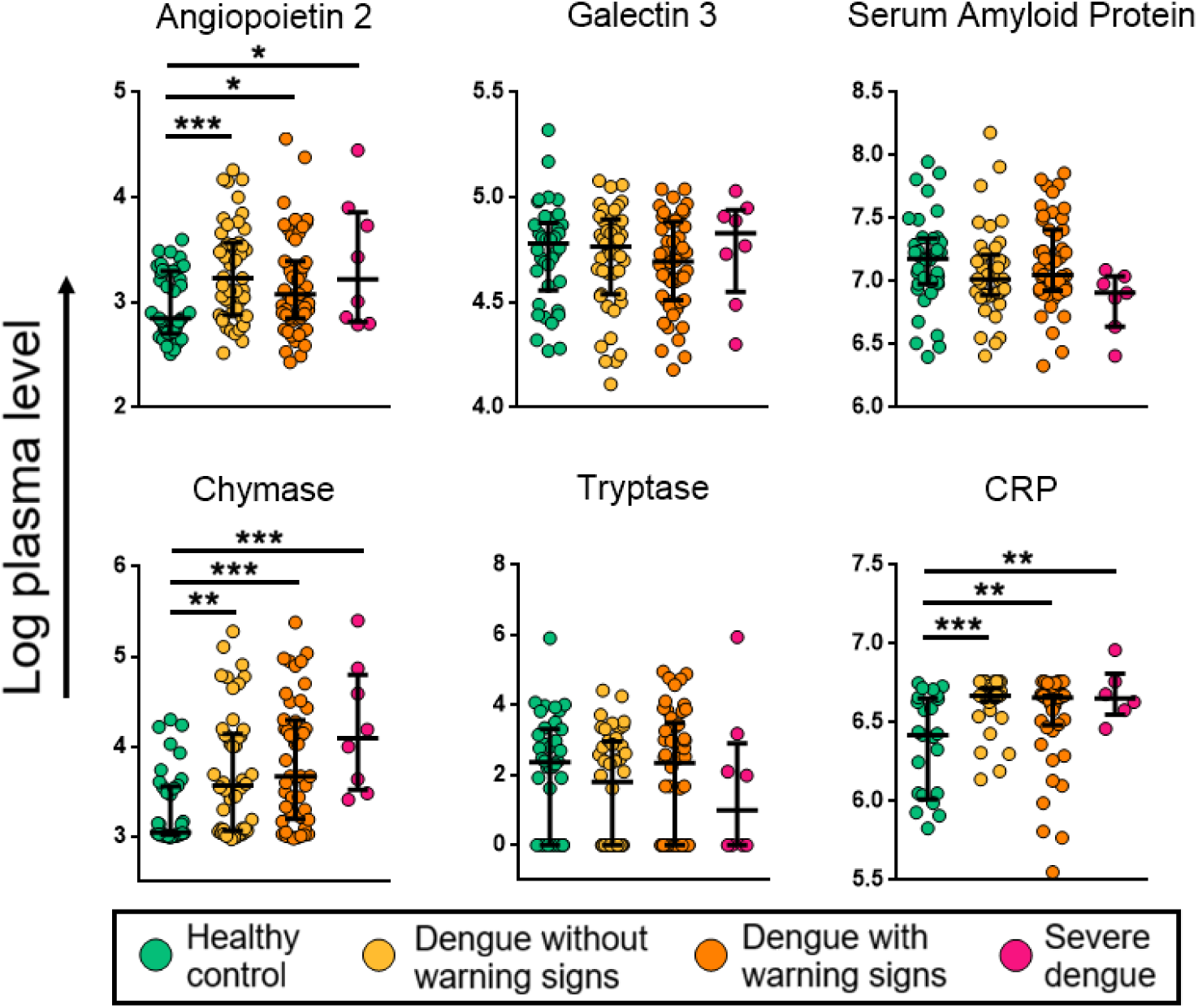
Comparison of serum levels of organ dysfunction analytes in dengue without warning signs, dengue with warning signs and severe dengue compared with healthy controls. Levels of biomarkers were compared across the four study groups by Kruskal–Wallis test. Post hoc Mann– Whitney U-tests were subsequently performed for those complement biomarkers with a Kruskal–Wallis P-value of <0.05. P-value <0.05 (significant); *, <0.05; **, <0.01; ***, <0.001, ****, <0.0001.

### Patients with dengue with warning signs displayed a relative systemic abundance of mediator’s suggestive of inflammation and hypercytokinemia

We next measured MIF levels (**Figure 2**) across the cohorts, which were significantly elevated among dengue patients as compared to healthy controls, with the highest increase detected in the DWS+ patients (P<0.01). Besides, TNF-α and IL-1β were also found to be elevated among dengue patients compared to healthy controls (**Figure 2**). Of note, TNF-α levels were significantly elevated among DWS+ (P<0.0001) as well as SD (P<0.01) patients but IL-1β showed a non-significant elevation across the group. Taken together, the findings indicate heighten inflammatory response consistent with acute inflammation and cytokine storm in clinical dengue infection.

### CRP levels showed significant increase whereas SAP and galectin 3 concentrations were not markedly different across the case-control cohorts

Having measured the levels of endogenous pyrogens TNF-α and IL-1β that are activators of production of acute-phase reactants by the liver, next, we determined the relative levels of mediators associated with hepatic injury and acute-phase reactants (often released by the liver) such as CRP, SAA and galectin 3. Our findings demonstrated that the levels of CRP (**Figure 3**) were significantly elevated in the dengue cohort as compared to healthy controls, with higher expression observed in the DWS+ (P<0.01) cohort and SD (P<0.01) than DWS– (P<0.001) and healthy while the levels of serum amyloid protein (**Figure 3**) and galectin 3 (**Figure 3**) did not show any significant alterations across the different case-control cohorts.

### Serum chymase levels were significantly elevated indicative of potential activation of mast cells in clinical dengue disease

The levels of chymase and tryptase, key markers of mast cell activation, were next assessed. Systemic chymase concentrations were significantly elevated both in the SD (P<0.001) as well as the DWS+ (P<0.001) cohorts compared to the DWS-(P<0.01) and healthy control groups. In contrast, serum tryptase levels (**Figure 3**) were varied with modest elevation observed among dengue patients but demonstrated a non-significant difference across the group. These findings suggest activation of mast cells during clinical dengue infection.

### Heightened levels of mediators associated with tissue repair and counter regulatory responses were evident in clinical dengue infection

We next examined levels of mediators involved in tissue repair and counter regulatory responses in clinical dengue infection. HGF levels (**Figure 2**) were modestly increased in dengue patients compared with controls, with slightly higher levels in severe dengue although there was no significant difference between the groups. In contrast, IL-1Ra (**Figure 2**) concentrations were significantly increased across the dengue groups, particularly in SD (P<0.01) and DWS+ (P<0.0001). These findings indicate activation of pathways associated with tissue repair and counter-regulatory responses in dengue infection. Similarly, IL-10 levels did not show significant variation between the cohorts.

### Broad elevation of inflammatory, endothelial, and mast cell mediators was observed in dengue infection across both primary and secondary cohorts

Next, we sought to compare the analytes between the primary and secondary dengue cohorts. Most of the analytes followed patterns similar to the earlier cohort stratification, albeit with a few notable changes. Of the endothelial biomarkers, CXCL10, TRAIL and Ang-2 showed significant differential expression across the cohorts, while CCL2 expression remained non-significant across the groups. CXCL10 expression was elevated in both primary (P<0.0001) and secondary dengue (p<0.0001) compared to the healthy controls. Ang-2 was significantly higher in primary dengue compared to healthy controls (P<0.001), and TRAIL levels were increased in both primary (P<0.05) and secondary dengue (P<0.05) as compared to healthy controls.

MIF, TNF-α, and IL-1β showed significant alterations across the cohorts, with MIF showing a significantly higher expression in primary dengue than in healthy controls (P<0.01), while TNF-α and IL-1β levels were significantly higher in both primary (TNF-α, P<0.0001; IL-1β, P<0.05) and secondary dengue (TNF-α, P<0.001; IL-1β, P<0.05) cohorts. SAP and galectin 3 showed non-significant alteration between the cohorts, while the expression of CRP was higher in primary (P<0.001) and secondary dengue (P<0.001) relative to the healthy controls. The levels of IL-1Ra were significantly increased in primary (P<0.0001) and secondary (P< 0.01) dengue. Likewise, the expression of mast cell-derived analytes also showed a similar pattern as in the earlier cohort stratifications as DWS+ and DWS-. Chymase levels were higher across both primary (P<0.01) and secondary dengue (P<0.0001) than in healthy controls. Moreover, chymase levels in secondary dengue wass significantly higher than in primary dengue (P<0.01), while tryptase concentrations remained non-significant across the cohorts **(Figures 4** and **5)**.

**Figure 4.**
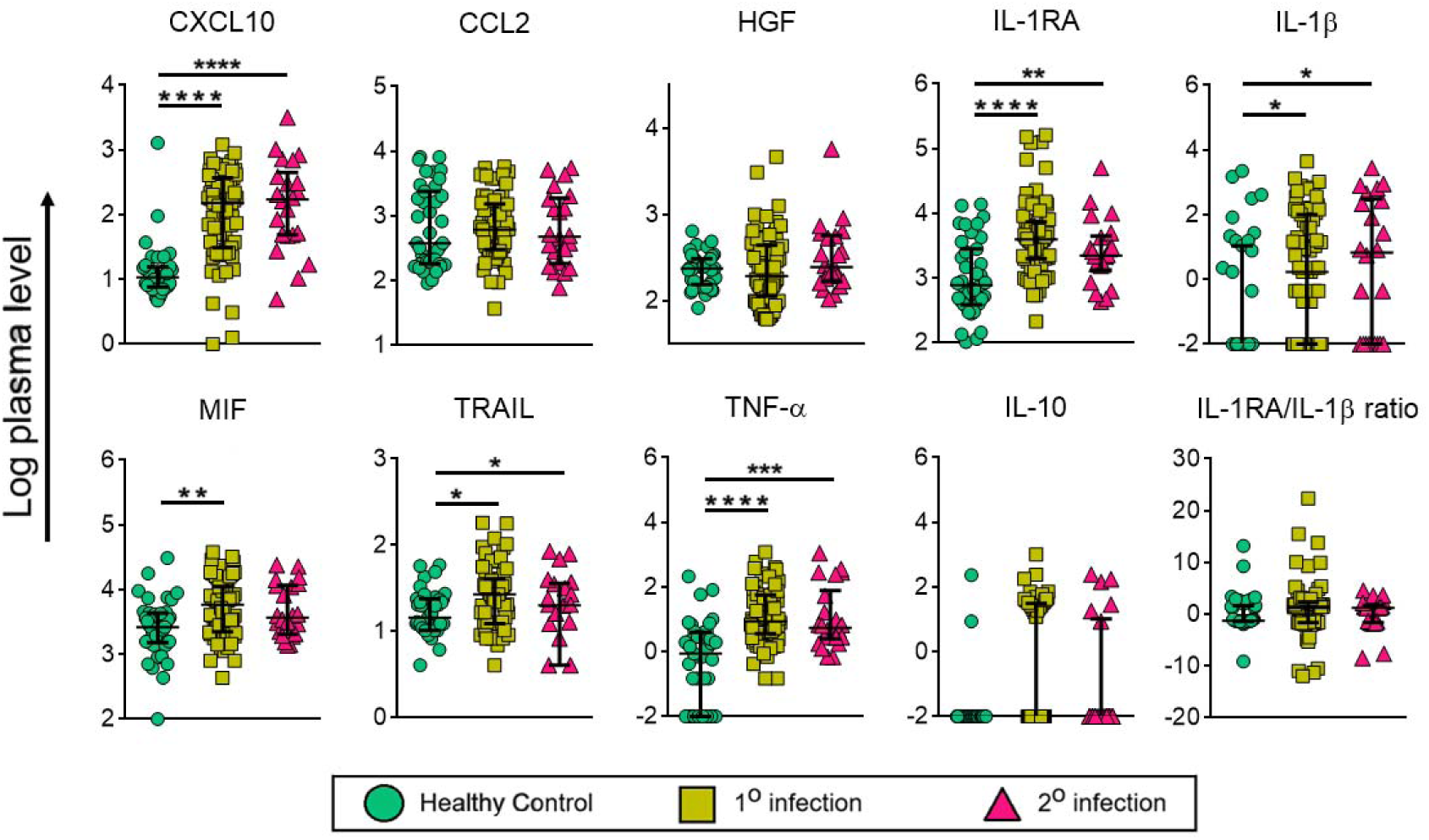
Comparison of serum levels of cytokines analytes in primary and secondary dengue with healthy controls. Levels of biomarkers were compared across the three study groups by Kruskal–Wallis test. Post hoc Mann– Whitney U-tests were subsequently performed for those complement biomarkers with a Kruskal–Wallis P-value of <0.05. P-value <0.05 (significant); *, <0.05; **, <0.01; ***, <0.001, ****, <0.0001.

**Figure 5.**
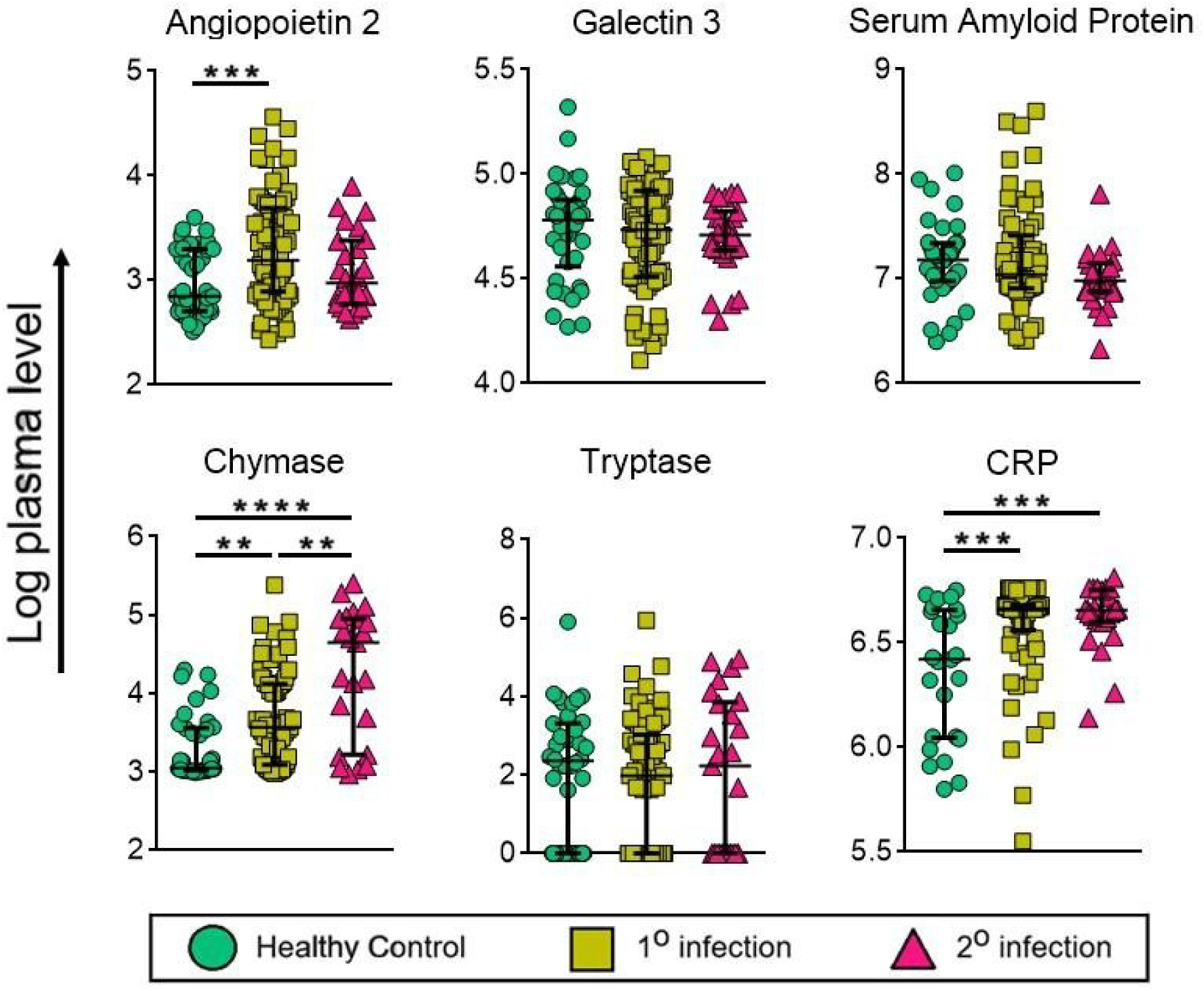
Comparison of serum levels of organ dysfunction analytes in primary and secondary dengue with healthy controls. Levels of biomarkers were compared across the three study groups by Kruskal–Wallis test. Post hoc Mann– Whitney U-tests were subsequently performed for those complement biomarkers with a Kruskal–Wallis P-value of <0.05. P-value <0.05 (significant); *, <0.05; **, <0.01; ***, <0.001, ****, <0.0001.

### Regression analysis revealed a strong positive association between disease severity and dengue progression

Next, we sought to perform a logistic regression analysis involving the different determinants of dengue severity, especially platelet counts and mediators and biomarkers of endothelial dysfunction. Initially, a logistic regression analysis was performed univariately to identify biosignatures that were significantly associated with dengue severity, platelet count, Ang-2, galectin 3 and SAP (**Figure 6A**). The mediators/biomarkers that showed a significant association in the univariate were regarded as candidate predictors and were further included for analysis by a multivariate regression. Our findings showed that Ang-2 demonstrated a strong positive correlation between severity and several disease progression indicators, such as markers associated with thrombocytopenia and hepatic dysfunction i.e. liver enzyme and bilirubin levels. Galectin 3 showed limited association with most clinical parameters, while SAP demonstrated modest positive association with inflammatory markers but showed minimal association with classical laboratory indicators of organ dysfunction (**Figure 6A**). Of the predictors of disease severity, ALT and GGT were positively associated with severity, while RDW showed a negative association. While, erythrocyte counts showed a negative association with platelet levels, cholesterol levels were positively associated with platelet counts (**Figure 6B**).

**Figure 6.**
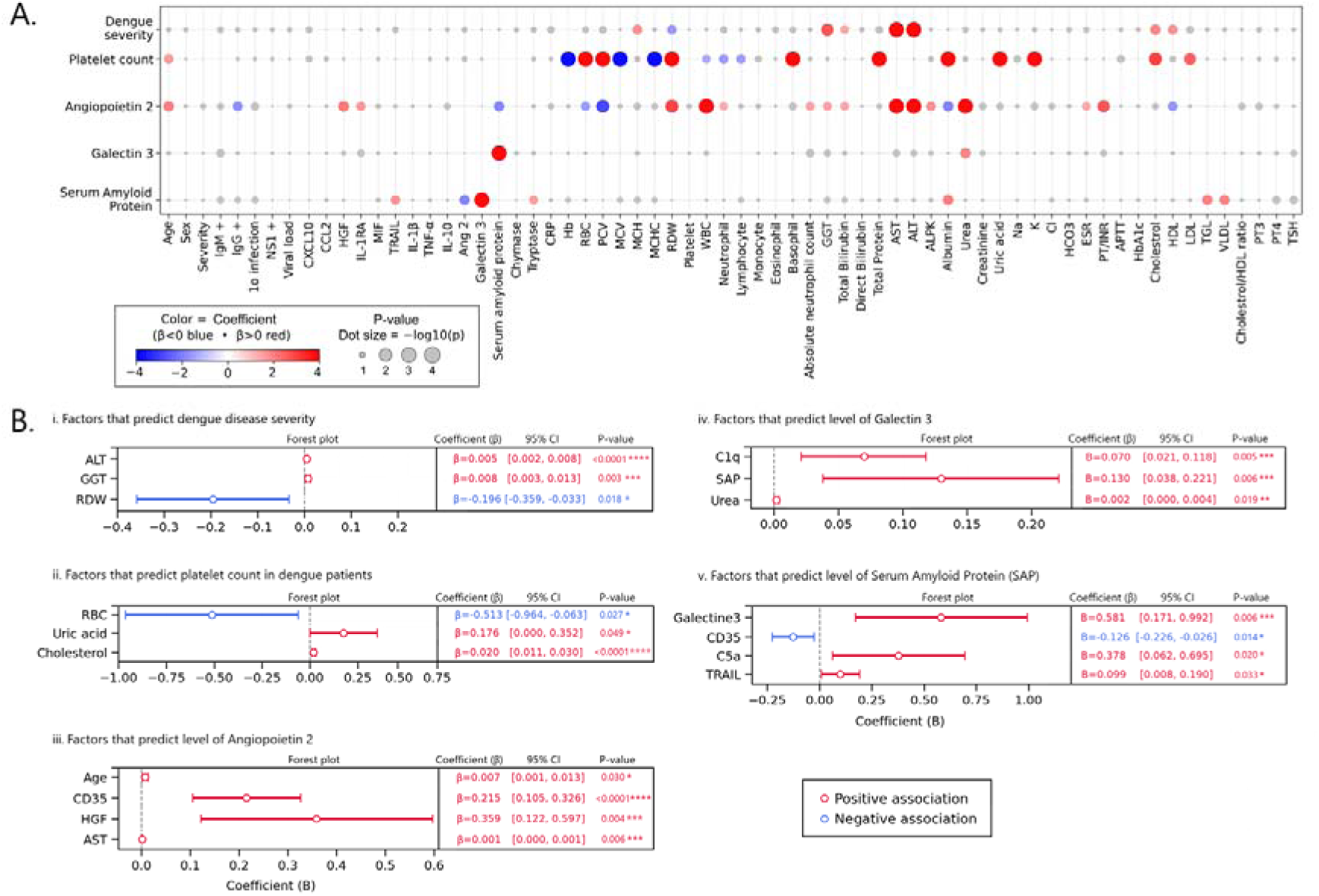
Factors associated with dengue severity, platelet counts and markers of endothelial dysfunction. (A) Univariate logistic regression analysis of parameters associated with dengue severity, platelet count, vascular endothelial dysfunction markers. The logistic regression analysis was performed univariately to identify biomarkers that were significantly associated with dengue severity, platelet count, Ang-2, galectin 3 and SAP (showed in heatmap above). The biomarkers that showed significant association in the univariate were regarded as candidate predictors and were included in multivariate regression analysis. The odds ratio (OR) and 95% confidence interval (CI) were estimated. (B) Forest plot for multivariate analyses – The candidate predictors were then included in a stepwise multivariate regression, and variables, for which p values were <0.05, were considered as independent predictors. *, **, ***, and **** represent P values <0.05, <0.01, <0.001, and <0.0001, respectively.

## Discussion

Our cross-sectional case-control study demonstrated that dengue severity is driven by coordinated activation of endothelial, inflammatory, mast cell-derived and counter-regulatory pathways that collectively promote vascular permeability and organ dysfunction. These findings highlight the complex interplay between immune activation, endothelial injury, and regulatory response in dengue pathogenesis. Among endothelial biomarkers, Ang-2 demonstrated a clear severity-dependent increase, with the highest levels observed among patients with warning signs and severe dengue (Mapalagamage et al., 2020). Ang-2 is a key antagonist ligand of the Tie2 pathway, which competitively inhibits Ang-1 binding. It is secreted in diseased vasculature and is implicated with angiogenesis and vascular stability (Zhang et al., 2019). It promotes endothelial destabilization and vascular leakage, and is also a known predictor of mortality in critically ill individuals (Mariko et al., 2019). In primary dengue, Ang-2 increase may reflect early endothelial activation via exocytosis of Weibel-Palade bodies. In primary dengue, the response is contingent upon naïve immunity without ADE-mediated Fc receptor, unlike severe secondary dengue, where Ang-2 emerges from viral-endothelial interactions or pro-inflammatory signals (Michels et al., 2012).

The surge in CXCL10 levels observed in DWS+ and severe dengue appears to hint its role in endothelial activation (Lachova et al., 2017). Further, increased CXCL10 could be linked to heightened increased dengue severity owing to exaggerated inflammation (Nor et al., 2018; Yong et al., 2017). Together with Ang-2 expression, this axis might be representative of dengue-associated inflammation and destabilisation of the endothelium entailing plasma leakage. Similarly, relative abundance of CXCL10 in primary and secondary dengue suggests an enhanced IFN-mediated antiviral responses and immune cell recruitment (Calderón-Peláez et al., 2022; Nwe et al., 2022).

Increased CCL2 is primarily indicative of monocyte recruitment rather than an indication of dengue disease severity (de-Oliveira-Pinto et al., 2012). In contrast, increased TRAIL levels in dengue indicates its role in immune mediated cytotoxic activity and endothelial stress, although its precise role in vascular dysfunction needs to be explored further. Functional studies indicate that TRAIL could be antiviral in DENV infection (Gandini et al., 2017; Limonta et al., 2014). The elevation of the TRAIL in the DWS+ and SD also aligns with immune regulation observed by others (Becerra et al., 2009).

Apart from endothelial dysfunction, elevated TNF-α and MIF further indicates the amplification of pro-inflammatory cascades in dengue. TNF-α is a well-established mediator of vascular leakage and activation, while MIF sustains inflammatory responses by counteracting glucocorticoid-mediated suppression. Hence, their elevation supports a hyperinflammatory milieu contributing to endothelial dysfunction. IL-1Ra was also markedly increased in dengue, consistent with activation of a compensatory anti-inflammatory response, whereas IL-1β showed variable expression without significant differences across the cohorts. This imbalance between pro-inflammatory response and counter-inflammatory pathway might be a reflection of immune dysregulation rather than a mere cytokine storm (de-Oliveira-Pinto et al., 2012; Imad et al., 2020; Tramontini Gomes de Sousa Cardozo et al., 2017). Unlike in our cohort based on dengue severity, IL-1β displayed significant differences when comparing primary and secondary dengue, with higher levels in secondary infection correlating with fever onset and vascular stress (de la Cruz Hernández et al., 2016).

CRP levels were markedly increased in dengue patients, particularly in DWS+ and severe dengue, indicating an amplified acute-phase response with hepatic involvement (Gregorova et al., 2025; Rao et al., 2020). In contrast, the SAP, galectin-3 and HGF levels remaining unchanged, suggests a selective rather than a global activation of the acute phase and growth factor response (Becquart et al., 2010; Fragnoud et al., 2015; Liu et al., 2016). The significant elevation in CRP in both primary and secondary dengue further reflects strong acute systemic inflammatory responses (Vuong et al., 2020).

The robust severity-dependent elevation of chymase in dengue patients, especially in DWS+ and severe dengue patients, support its role in mast cell activation (Rathore et al., 2020; Tissera et al., 2017). Chymase is known to be involved in disruption of endothelial barriers, matrix remodelling and vascular permeability (Rathore et al., 2020; Tissera et al., 2017). The parallel elevation of chymase as well as Ang-2-CXCL10 axis supports a mast cell-endothelial interaction contributing to plasma leakage. In contrast, the low levels of tryptase, another mast cell-derived mediator is suggestive of a selective mast cell mediator release rather than global degranulation in dengue severity (Sherif et al., 2020; Teo et al., 2024).

The strong positive association of Ang-2 with liver enzymes and thrombocytopenia markers further reinforces the earlier observation of a strong link between the endothelial injury and systemic organ involvement (McBride et al., 2024). Regression analysis identified certain factors associated with inflammation. Age, HGF, and AST were significantly associated with Ang-2 levels, suggesting a potential link between hepatic functions and endothelial dysfunction. SAP and urea were associated with galectin-3, suggesting interactions between the acute-phase reactants and metabolic factors. In addition, SAP levels were significantly associated with galectin-3, CD163, and TRAIL, highlighting their likely role in macrophage stimulation, complement activation and immune signalling (Anshad et al., 2026).

Our study has certain limitations that need to be acknowledged, such as the non-addressing of potential confounding factors including co-morbidities, as well as other flavivirus infections. Also, the plaque reduction neutralisation test for confirming DENV infection was not performed due to sample volume constraints. In addition, the cross-sectional nature of the study fails to establish a spatio-temporal relationship between the analytes across the varying severity of the disease.

In conclusion, our study demonstrates that dengue severity is driven by coordinated endothelial dysfunction, heightened inflammatory responses, mast cell-derived mediators, and counter-regulatory immune pathways that collectively contribute to vascular leakage and organ injury. Of all the analytes measured, Ang-2, CXCL10, TRAIL, CRP, MIF, IL-1Ra, TNF-α, chymase, and CRP were strongly associated with disease severity, highlighting the central role of endothelial activation and IFN-driven inflammation in dengue pathogenesis. Correlation analysis further indicated endothelial injury, as Ang-2 showed a strong positive association with thrombocytopenia and hepatic injury markers. Elevated chymase levels suggest mast cell–driven vascular damage and potential mast cell–endothelial interactions contributing to capillary leakage, while increased CRP reflects both acute-phase responses as well as hepatic involvement. Overall, these findings identify several biomarkers linked to dengue severity. Addressing these gaps would require future studies that integrate comprehensive longitudinal sampling with expanded immunological profiling.

## Data Availability

All data produced in the present work are contained in the manuscript.

## Acknowledgements

The authors thank all the national and international members of the Infectious Diseases Society of India (IDSI), Chennai, for extending insightful discussions as well as logistic support. The authors would also like to acknowledge the SRMDBT facility, SRM Institute of Science and Technology, Chennai.

## Author contributions

A.R.A, M.A, S.S, A.M, M.L, Y.K, P.B, S.R, and E.M.S. designed the study and were responsible for conceptualisation and data curation. A.R.A, M.A, M.L., A.M, and E.M.S conducted the analysis and were responsible for methodology, formal analysis, validation, and visualisation. A.R.A, S.S., P.B., and E.M.S. wrote the first draft of the manuscript. All authors reviewed the manuscript.

## Funding

A.R.A is funded by NFOBC-NBCFDC of the Ministry of Social Justice, Government of India, as Senior Research Fellowship. The authors also acknowledge funding support received from M.L. through 2021-02703, the Swedish Research Council (https://www.vr.se/english.html), The Joanna Cocozza Foundation for Children’s Medical Research, and Linköping University Hospital Research, CALF. The funders of the study had no role in the study design, data collection, data analysis, data interpretation, or writing of the report.

## Conflict of interest

The author(s) declared that this work was conducted in the absence of any commercial or financial relationships that could be construed as a potential conflict of interest.

## Notes

### Competing Interest Statement

The authors have declared no competing interest.

### Author Declarations

Ethics committee/IRB of SMCH, Chennai (Ref. No. 114/03/2024/Faculty/SRB/SMCH) and TMC, Theni (Ref. No. 2300/IEC/2024-26) gave ethical approval for this work.

## References

1. Anshad, A. R., Saravanan, S., Murugesan, A., Vighnesh, R., Raju, S., Kannan, R., Yong, Y. K., Larsson, M., & Shankar, E. M. (2026). Aberrant systemic acute-phase complement responses in conjunction with soluble CR1 attribute to varying grades of dengue disease severity. Frontiers in Immunology, 16. 10.3389/fimmu.2025.1731011

2. Becerra, A., Warke, R. V., Martin, K., Xhaja, K., de Bosch, N., Rothman, A. L., & Bosch, I. (2009). Gene expression profiling of dengue infected human primary cells identifies secreted mediators in vivo. Journal of Medical Virology, 81(8), 1403–1411. 10.1002/jmv.21538

3. Becquart, P., Wauquier, N., Nkoghe, D., Ndjoyi-Mbiguino, A., Padilla, C., Souris, M., & Leroy, E. M. (2010). Acute dengue virus 2 infection in Gabonese patients is associated with an early innate immune response, including strong interferon alpha production. BMC Infectious Diseases, 10(1), 356. 10.1186/1471-2334-10-356

4. Brasier, A. R., Ju, H., Garcia, J., Spratt, H. M., Victor, S. S., Forshey, B. M., Halsey, E. S., Comach, G., Sierra, G., Blair, P. J., Rocha, C., Morrison, A. C., Scott, T. W., Bazan, I., Kochel, T. J., &. (2012). A Three-Component Biomarker Panel for Prediction of Dengue Hemorrhagic Fever. The American Society of Tropical Medicine and Hygiene, 86(2), 341–348. 10.4269/ajtmh.2012.11-0469

5. Calderón-Peláez, M.-A., Coronel-Ruiz, C., Castellanos, J. E., & Velandia-Romero, M. L. (2022). Endothelial Dysfunction, HMGB1, and Dengue: An Enigma to Solve. Viruses, 14(8), 1765. 10.3390/v14081765

6. Castilho, B. M., Silva, M. T., Freitas, A. R. R., Fulone, I., & Lopes, L. C. (2020). Factors associated with thrombocytopenia in patients with dengue fever: a retrospective cohort study. BMJ Open, 10(9), e035120. 10.1136/bmjopen-2019-035120

7. Cipitelli, M. da C., Paiva, I. A., Badolato-Corrêa, J., Marinho, C. F., Fiestas Solórzano, V. E., da Costa Faria, N. R., de Azeredo, E. L., de Souza, L. J., da Cunha, R. V., & De-Oliveira-Pinto, L. M. (2022). Subsets of Cytokines and Chemokines from DENV-4-Infected Patients Could Regulate the Endothelial Integrity of Cultured Microvascular Endothelial Cells. Pathogens, 11(5), 509. 10.3390/pathogens11050509

8. Conroy, A. L., Gélvez, M., Hawkes, M., Rajwans, N., Tran, V., Liles, W. C., Villar-Centeno, L. A., & Kain, K. C. (2015). Host biomarkers are associated with progression to dengue haemorrhagic fever: a nested case-control study. International Journal of Infectious Diseases, 40, 45–53. 10.1016/j.ijid.2015.07.027

9. Dalrymple, N. A., & Mackow, E. R. (2012). Endothelial Cells Elicit Immune-Enhancing Responses to Dengue Virus Infection. Journal of Virology, 86(12), 6408–6415. 10.1128/JVI.00213-12

10. de-Oliveira-Pinto, L. M., Gandini, M., Freitas, L. P., Siqueira, M. M., Marinho, C. F., Setúbal, S., Kubelka, C. F., Cruz, O. G., & Oliveira, S. A. de. (2012). Profile of circulating levels of IL-1Ra, CXCL10/IP-10, CCL4/MIP-1β and CCL2/MCP-1 in dengue fever and parvovirosis. Memórias Do Instituto Oswaldo Cruz, 107(1), 48–56. 10.1590/S0074-02762012000100007

11. de la Cruz Hernández, S. I., Puerta-Guardo, H. N., Flores Aguilar, H., González Mateos, S., López Martinez, I., Ortiz-Navarrete, V., Ludert, J. E., & del Angel, R. M. (2016). Primary dengue virus infections induce differential cytokine production in Mexican patients. Memórias Do Instituto Oswaldo Cruz, 111(3), 161–167. 10.1590/0074-02760150359

12. Fragnoud, R., Flamand, M., Reynier, F., Buchy, P., Duong, V., Pachot, A., Paranhos-Baccala, G., & Bedin, F. (2015). Differential proteomic analysis of virus-enriched fractions obtained from plasma pools of patients with dengue fever or severe dengue. BMC Infectious Diseases, 15(1), 518. 10.1186/s12879-015-1271-7

13. Gandini, M., Petitinga-Paiva, F., Marinho, C. F., Correa, G., De Oliveira-Pinto, L. M., Souza, L. J. de, Cunha, R. V., Kubelka, C. F., & Azeredo, E. L. de. (2017). Dengue Virus Induces NK Cell Activation through TRAIL Expression during Infection. Mediators of Inflammation, 2017, 1–10. 10.1155/2017/5649214

14. Giang, N. T., van Tong, H., Quyet, D., Hoan, N. X., Nghia, T. H., Nam, N. M., Hung, H. V., Anh, D. T., Van Mao, C., Son, H. A., Meyer, C. G., Velavan, T. P., & Toan, N. L. (2020). Complement protein levels and MBL2 polymorphisms are associated with dengue and disease severity. Scientific Reports, 10(1), 14923. 10.1038/s41598-020-71947-2

15. Goswami, R. P., Mukherjee, A., Biswas, T., Karmakar, P. S., & Ghosh, A. (2012). Two cases of dengue meningitis: A rare first presentation. Journal of Infection in Developing Countries, 6(2), 208–211. 10.3855/jidc.2241

16. Gregorova, M., Santopaolo, M., Garner, L. C., Hayati, R. F., Diamond, D., Ramamurthy, N., Tran, V. T., Nguyen, N. M., Heesom, K. J., Nguyen, V. L., Jones, E., Nsubuga, M., Luscombe, C., Vo, H. T. M., Ho, C. Q., Nguyen, C. T. X., Dong, T. T. H., Huynh, D. T. Le, Cao, T. T., … Rivino, L. (2025). Early NK-cell and T-cell dysfunction marks progression to severe dengue in patients with obesity and healthy weight. Nature Communications, 16(1), 5569. 10.1038/s41467-025-60941-9

17. Gurugama, P., Jayarajah, U., Wanigasuriya, K., Wijewickrama, A., Perera, J., & Seneviratne, S. L. (2018). Renal manifestations of dengue virus infections. Journal of Clinical Virology, 101, 1–6. 10.1016/j.jcv.2018.01.001

18. Halstead, S., & Wilder-Smith, A. (2019). Severe dengue in travellers: Pathogenesis, risk and clinical management. Journal of Travel Medicine, 26(7), 1–15. 10.1093/jtm/taz062

19. Imad, H. A., Phumratanaprapin, W., Phonrat, B., Chotivanich, K., Charunwatthana, P., Muangnoicharoen, S., Khusmith, S., Tantawichien, T., Phadungsombat, J., Nakayama, E., Konishi, E., & Shioda, T. (2020). Cytokine Expression in Dengue Fever and Dengue Hemorrhagic Fever Patients with Bleeding and Severe Hepatitis. The American Journal of Tropical Medicine and Hygiene, 102(5), 943–950. 10.4269/ajtmh.19-0487

20. Keshav, L. B., K, A., Malhotra, K., & Shetty, S. (2024). Lung Manifestation of Dengue Fever: A Retrospective Study. Cureus. 10.7759/cureus.60655

21. Khanam, A., Gutiérrez-Barbosa, H., Lyke, K. E., & Chua, J. V. (2022). Immune-Mediated Pathogenesis in Dengue Virus Infection. Viruses, 14(11), 2575. 10.3390/v14112575

22. Lachová, V., Škorvanová, L., Svetlíková, D., Turianová, L., Kostrábová, A., & Betáková, T. (2017). Comparison of transcriptional profiles of interferons, CXCL10 and RIG-1 in influenza infected A549 cells stimulated with exogenous interferons. Acta Virologica, 61(02), 183–190. 10.4149/av_2017_02_07

23. Lai, Y.-C., Chao, C.-H., & Yeh, T.-M. (2020). Roles of Macrophage Migration Inhibitory Factor in Dengue Pathogenesis: From Pathogenic Factor to Therapeutic Target. Microorganisms, 8(6), 891. 10.3390/microorganisms8060891

24. Limonta, D., Torrentes□Carvalho, A., Marinho, C. F., de Azeredo, E. L., de Souza, L. J., Motta□Castro, A. R. C., da Cunha, R. V., Kubelka, C. F., Nogueira, R. M. R., & De□Oliveira□Pinto, L. M. (2014). Apoptotic mediators in patients with severe and non□severe dengue from Brazil. Journal of Medical Virology, 86(8), 1437–1447. 10.1002/jmv.23832

25. Liu, K.-T., Liu, Y.-H., Chen, Y.-H., Lin, C.-Y., Huang, C.-H., Yen, M.-C., & Kuo, P.-L. (2016). Serum Galectin-9 and Galectin-3-Binding Protein in Acute Dengue Virus Infection. International Journal of Molecular Sciences, 17(6), 832. 10.3390/ijms17060832

26. Mapalagamage, M., Handunnetti, S. M., Wickremasinghe, A. R., Premawansa, G., Thillainathan, S., Fernando, T., Kanapathippillai, K., De Silva, A. D., & Premawansa, S. (2020). High Levels of Serum Angiopoietin 2 and Angiopoietin 2/1 Ratio at the Critical Stage of Dengue Hemorrhagic Fever in Patients and Association with Clinical and Biochemical Parameters. Journal of Clinical Microbiology, 58(4). 10.1128/JCM.00436-19

27. Mariko, R., Darwin, E., Yanwirasti, Y., & Hadinegoro, S. R. (2019). The Difference of Angiopoietin-2 Levels between Dengue Hemorrhagic Fever Patients with Shock and without Shock. Open Access Macedonian Journal of Medical Sciences, 7(13), 2119–2122. 10.3889/oamjms.2019.569

28. McBride, A., Duyen, H. T. Le, Vuong, N. L., Tho, P. V., Tai, L. T. H., Phong, N. T., Ngoc, N. T., Yen, L. M., Nhat, P. T. H., Vi, T. T., Llewelyn, M. J., Thwaites, L., Hao, N. Van, & Yacoub, S. (2024). Endothelial and inflammatory pathophysiology in dengue shock: New insights from a prospective cohort study in Vietnam. PLOS Neglected Tropical Diseases, 18(3), e0012071. 10.1371/journal.pntd.0012071

29. Meena, A. A., Murugesan, A., Sopnajothi, S., Yong, Y. K., Ganesh, P. S., Vimali, I. J., Vignesh, R., Elanchezhiyan, M., Kannan, M., Dash, A. P., & Shankar, E. M. (2020). Increase of Plasma TNF-α Is Associated with Decreased Levels of Blood Platelets in Clinical Dengue Infection. Viral Immunology, 33(1), 54–60. 10.1089/vim.2019.0100

30. Mehmood, A., Afzal, M. W., Ahmad, M., Mufti, M., Malik, J., & Zaidi, S. M. J. (2023). Respiratory sequelae of dengue fever. Tropical Doctor, 53(2), 237–240. 10.1177/00494755221127355

31. Michels, M., van der Ven, A. J. A. M., Djamiatun, K., Fijnheer, R., de Groot, P. G., Griffioen, A. W., Sebastian, S., Faradz, S. M. H., & de Mast, Q. (2012). Imbalance of Angiopoietin-1 and Angiopoetin-2 in Severe Dengue and Relationship with Thrombocytopenia, Endothelial Activation, and Vascular Stability. The American Society of Tropical Medicine and Hygiene, 87(5), 943–946. 10.4269/ajtmh.2012.12-0020

32. Mishra, H., Ngai, M., Crowley, V. M., Tran, V., Painaga, M. S. S., Gaite, J. Y., Hamilton, P., Kain, K. C., & Hawkes, M. T. (2024). The Angiopoietin–Tie-2 Axis in Children and Young Adults with Dengue Virus Infection in the Philippines. American Journal of Tropical Medicine and Hygiene, 111(4), 887–896. 10.4269/ajtmh.24-0115

33. Mukherjee, S., Saha, B., & Tripathi, A. (2022). Clinical significance of differential serum-signatures for early prediction of severe dengue among Eastern Indian patients. Clinical and Experimental Immunology, 208(1), 72–82. 10.1093/cei/uxac018

34. Nanaware, N., Banerjee, A., Mullick Bagchi, S., Bagchi, P., & Mukherjee, A. (2021). Dengue Virus Infection: A Tale of Viral Exploitations and Host Responses. Viruses, 13(10), 1967. 10.3390/v13101967

35. Narvaez, F., Montenegro, C., Juarez, J. G., Zambrana, J. V., Gonzalez, K., Videa, E., Arguello, S., Barrios, F., Ojeda, S., Plazaola, M., Sanchez, N., Camprubí-Ferrer, D., Kuan, G., Paz Bailey, G., Harris, E., & Balmaseda, A. (2025). Dengue severity by serotype and immune status in 19 years of pediatric clinical studies in Nicaragua. PLOS Neglected Tropical Diseases, 19(1), e0012811. 10.1371/journal.pntd.0012811

36. Nor, F. M., Khamis, S. N., Wang, S. M., Mokhtar, M. A. M., Yuhana, M. Y., Hoh, B. P., & Zain, Z. M. (2018). Evaluation of CXCL10 as potential biomarker for early detection of severe dengue. International Journal of Infectious Diseases, 73, 395. 10.1016/j.ijid.2018.04.4310

37. Nwe, K. M., Ngwe Tun, M. M., Myat, T. W., Sheng Ng, C. F., Htun, M. M., Lin, H., Hom, N. S., Soe, A. M., Elong Ngono, A., Hamano, S., Morita, K., Thant, K. Z., Shresta, S., Thu, H. M., & Moi, M. L. (2022). Acute-phase Serum Cytokine Levels and Correlation with Clinical Outcomes in Children and Adults with Primary and Secondary Dengue Virus Infection in Myanmar between 2017 and 2019. Pathogens, 11(5), 558. 10.3390/pathogens11050558

38. Paz-Bailey, G., Adams, L. E., Deen, J., Anderson, K. B., & Katzelnick, L. C. (2024). Dengue. The Lancet, 403(10427), 667–682. 10.1016/S0140-6736(23)02576-X

39. Rao, R., Nayak, S., Pandey, A., & Kamath, S. (2020). Diagnostic performance of C-reactive protein level and its role as a potential biomarker of severe dengue in adults. Asian Pacific Journal of Tropical Medicine, 13(8), 358. 10.4103/1995-7645.289440

40. Rathore, A. P. S., Senanayake, M., Athapathu, A. S., Gunasena, S., Karunaratna, I., Leong, W. Y., Lim, T., Mantri, C. K., Wilder-Smith, A., & St. John, A. L. (2020). Serum chymase levels correlate with severe dengue warning signs and clinical fluid accumulation in hospitalized pediatric patients. Scientific Reports, 10(1), 11856. 10.1038/s41598-020-68844-z

41. Samanta, J. (2015). Dengue and its effects on liver. World Journal of Clinical Cases, 3(2), 125. 10.12998/wjcc.v3.i2.125

42. Sánchez, J. L., Salgado, D. M., Vega, M. R., Castro-Trujillo, S., & Narváez, C. F. (2024). Utility of the WHO dengue guidelines in pediatric immunological studies. Journal of Tropical Pediatrics, 70(4). 10.1093/tropej/fmae014

43. Sherif, N. A., Zayan, A. H., Elkady, A. H., Ghozy, S., Ahmed, A. R., Omran, E. S., Taha, E. A., Eldesoky, E. A., Ebied, A., Tieu, T., Maraie, N., Kamel, M. G., Ngo, H. T., Mattar, O. M., Hirayama, K., & Huy, N. T. (2020). Mast cell mediators in relation to dengue severity: A systematic review and meta□analysis. Reviews in Medical Virology, 30(1). 10.1002/rmv.2084

44. Teo, A., Le, C. T. T., Tan, T., Chia, P. Y., & Yeo, T. W. (2024). Febrile Phase Soluble Urokinase Plasminogen Activator Receptor and Olfactomedin 4 as Prognostic Biomarkers for Severe Dengue in Adults. Clinical Infectious Diseases, 78(3), 788–796. 10.1093/cid/ciad637

45. Tissera, H., Rathore, A. P. S., Leong, W. Y., Pike, B. L., Warkentien, T. E., Farouk, F. S., Syenina, A., Eong Ooi, E., Gubler, D. J., Wilder-Smith, A., & St. John, A. L. (2017). Chymase Level Is a Predictive Biomarker of Dengue Hemorrhagic Fever in Pediatric and Adult Patients. The Journal of Infectious Diseases, 216(9), 1112–1121. 10.1093/infdis/jix447

46. Tramontini Gomes de Sousa Cardozo, F., Baimukanova, G., Lanteri, M. C., Keating, S. M., Moraes Ferreira, F., Heitman, J., Pannuti, C. S., Pati, S., Romano, C. M., & Cerdeira Sabino, E. (2017). Serum from dengue virus-infected patients with and without plasma leakage differentially affects endothelial cells barrier function in vitro. PLOS ONE, 12(6), e0178820. 10.1371/journal.pone.0178820

47. Vaidya, A., & Salvi, S. P. (2023). Gastrointestinal and hepatic manifestations and outcome in dengue multi-organ dysfunction syndrome: data from a tertiary referral center. Infectious Diseases and Tropical Medicine, 9. 10.32113/idtm_20239_1181

48. Vuong, N. L., Le Duyen, H. T., Lam, P. K., Tam, D. T. H., Vinh Chau, N. Van, Van Kinh, N., Chanpheaktra, N., Lum, L. C. S., Pleités, E., Jones, N. K., Simmons, C. P., Rosenberger, K., Jaenisch, T., Halleux, C., Olliaro, P. L., Wills, B., & Yacoub, S. (2020). C-reactive protein as a potential biomarker for disease progression in dengue: a multi-country observational study. BMC Medicine, 18(1), 35. 10.1186/s12916-020-1496-1

49. Yong, Y. K., Tan, H. Y., Jen, S. H., Shankar, E. M., Natkunam, S. K., Sathar, J., Manikam, R., & Sekaran, S. D. (2017). Aberrant monocyte responses predict and characterize dengue virus infection in individuals with severe disease. Journal of Translational Medicine, 15(1), 121. 10.1186/s12967-017-1226-4

50. Zhang, Y., Kontos, C. D., Annex, B. H., & Popel, A. S. (2019). Angiopoietin-Tie Signaling Pathway in Endothelial Cells: A Computational Model. IScience, 20, 497–511. 10.1016/j.isci.2019.10.006

